# Are Mobile Phones part of the chain of transmission of SARS-CoV-2 in the hospital?

**DOI:** 10.1101/2020.11.02.20224519

**Authors:** Evelyn Patricia Sánchez Espinoza, Marina Cortes Farrel, Saidy Vasconez Nogueira, Anderson Vicente de Paula, Thais Guimarães, Lucy Santos Villas Boas, Marcelo Park, Cristina Carvalho da Silva, Ingra Morales, Lauro Vieira Perdigão Neto, Tania Regina Tozetto-Mendoza, Icaro Boszczowski, Ester Sabino, Maria Cássia Mendes-Correa, Anna Sara Shafferman Levin, Silvia Figueiredo Costa

**Affiliations:** Department of Infectious Diseases, Faculdade de Medicina, Universidade de São Paulo, São Paulo, Brazil; ^LIM52^ Virology Laboratory, Universidade de São Paulo, São Paulo, Brazil; Hospital das Clinicas, Faculdade de Medicina, Universidade de São Paulo, São Paulo, Brazil; Department of Infectious Diseases, Faculdade de Medicina, Universidade de São Paulo. Brazil; Department of Infectious Diseases, Faculdade de Medicina, Universidade de São Paulo, Brazil; Department of Infectious Diseases, Faculdade de Medicina, Universidade de São Paulo; Department of Infectious Diseases, Instituto de Medicina Tropical, Universidade de São Paulo, Brazil

**Keywords:** SARS-CoV-2 hospital cross-contamination, Healthcare workers mobile phones, SARS-CoV-2 fomites

## Abstract

SARS-CoV-2 cross-transmission has become an concern in hospitals. We investigate healthcare workers(HCWs) knowledge about SARS-CoV-2 cross-transmission and conceptions whether the virus can remain on HCWs mobile phones(MPs) and be part of the chain of transmission.

A cross-sectional study was conducted at a COVID-19 Intensive Care Unit of a teaching-hospital. Fifty-one MPs were swabbed and a questionnaire about hand hygiene and MP use and disinfection was applied after an educational campaign. Although most of HCWs believed on the importance of cross-transmission and increased hand hygiene adhesion and MP disinfection during the pandemic, SARS-CoV-2 RNA was detected in two MPs(culture of the samples was negative).

Implementation of official hospital policies to guide HCWs regarding disinfection and care of personal MP are needed.

## Introduction

Mobile phones(MPs) have become a working-tool. However, there are no official policies from the Centres for Disease Control and Prevention(CDC) about its disinfection in healthcare facilities. Permanence of SARS-CoV-2 in surfaces of hospital environment has been described^1^ raising the concern about cross-transmission^2^. Even though SARS-CoV-2 has been found in MPs of patients with COVID-19^3^, they have not been portrayed as source of transmission in the hospital.

## Material and methods

This is a cross-sectional study performed at an adult Intensive Care Unit(ICU) of a teaching hospital in Sao Paulo, Brazil. The ICU has 11 separated patient-rooms.

Healthcare workers(HCWs) use scrubs, N95-respirators and surgical-cap inside of the unit and add a surgical-gown, face shield and gloves when entering a patient’s room.

An educational campaign about cross-transmission and disinfection of MPs was performed at the beginning of the pandemic. Informative posters were left in the unit that had a QR-code with access to a video of the campaign advising to use 70% alcohol swab on MPs and to place a screen protector to protect the MP oleophobic coating; and avoid the use of MPs during patient care and in the restroom.

Ten days after the campaign we collected samples from HCW’s MPs. Herewith an electronic questionnaire was applied. It included queried conceptions about hand hygiene and MP care.

The MPs were swabbed using a nylon FLOQ Swab™(Copan Italia SPA-Italy) beginning with the front-screen and without taking its cases off. Once sampled, swabs were placed in universal transport medium(UTM™ Copan Italia SPA-Italy) stored at -80ºC and submitted to SARS-CoV-2 RT-PCR. Positive RT-PCR samples were submitted to viral culture.

The extraction of nucleic acids was performed using QIAmp viral RNA mini kit(QIAGEN-USA). For the RT-PCR assay, the commercial RealStar®SARS-CoV-2 Kit1.0(Altona-Diagnostics-Germany) that detects the presence of SARS-CoV-2 through amplification of the S-gene and E-gene was used. The amplification was done using the Roche Light Cycler® 96System(USA). According to the manufacturer, the sample is considered positive when one of the target genes is detected.

Culture was performed inoculating Vero cells (ATCC® CCL-81™), as previously described ^45^, in Dulbecco minimal essential medium supplemented with fetal bovine serum(5%), antibiotics and antimycotics(Cultilab, Campinas, São Paulo, www.cultilb.com.br) at 37°C in an atmosphere with 5%CO2. Every day the cells were examined for cytopathic effect(CE), new RT-PCR of the samples and blind passage of culture supernatant was performed on the third, seven and fourteen day of culture.

## Results

Fifty-one of the fifty-three HCWs working in the unit participated on the survey and responded the questionnaire. Nine(18%) had covered their MP with kitchen-plastic film in an attempt to facilitate disinfection. Eleven(16%) did not recall the educational campaign and three(6%) answered that it did not change their behaviour. Only four(8%) did not believe that the virus could remain on MPs and one(4%) did not believe that the virus could remain on the hands; 98% referred washing their hands more since the pandemic(Table1).

**Table 1.** Detection of SARS-CoV-2 by real-time polymerase chain reaction(RT-PCR) on mobile phones (MPs), and healthcare workers’ ideas on hand hygiene and mobile phone use during the pandemic

| RT-PCR result / Answer to questionnaire | Attending physician<br>(n=7) | Cleaning staff<br>(n=5) | Nurse<br>(n=7) | Nursing assistant<br>(n=17) | Physiotherapist<br>(n=5) | Resident/<br>Fellow<br>(n=10) | Total<br>(n=51) |
| --- | --- | --- | --- | --- | --- | --- | --- |
| Positive RT-PCR on Mobile phones | 0 | 0 | 1(14%) | 1(6%) | 0 | 0 | 3(6%) |
| Believes SARS-CoV-2 can stay on hands | 7(100%) | 4(80%) | 7(100%) | 16(94%) | 5(100%) | 10(100%) | 49(96%) |
| Increased hand-hygiene after COVID-19 | 6(86%) | 5(100%) | 7(100%) | 17(100%) | 5(100%) | 10(100%) | 50(98%) |
| Believes SARS-CoV-2 can remain on MPs | 7(100%) | 4(80%) | 7(100%) | 14(82%) | 5(100%) | 10(100%) | 47(92%) |
| Increased cleaning of MP after COVID-19 | 6(86%) | 5(100%) | 7(100%) | 16(94%) | 5(100%) | 9(90%) | 48(94%) |

Twenty-two COVID-19 patients were hospitalized in the unit, eleven(50%) had positive SARS-CoV-2 RT-PCR and twenty(91%) had a lung computed-tomography suggestive of COVID-19.

Fifty-one MP swabs were collected, two were positive by RT-PCR(4%), with C_t_ values of 34 and 36, both detected the E-gene.

The two RT-PCR positive samples were isolated for viral culture. The sample with C_t_ 34 showed CE on the third day, subsequent RT-PCR of this isolate was negative. The isolate with C_t_ 36 did not show CE, subsequent RT-PCR of this isolate was negative as well. The supernatant of both cultures were monitored for 14 days without observing any other CE afterwards.

## Discussion

In this study, although most of HCWs believed on the importance of cross-transmission and increased the adhesion to hand hygiene and MP disinfection during the pandemic, we identified SARS-CoV-2 on MPs. Our findings suggest the need of a universal policy in the infection-control guidelines on how to care for electronic devices in the hospital.

Little is known about virus on MPs or its potential for cross-contamination. A study of MPs from HCWs of a Paediatric unit^6^ found virus RNA in 38.5% of the cases(42/109); predominantly Norovirus(n=39).

Two samples from a CPAP helmet used by COVID-19 patients, were positive by the RT-PCR[1] from patients who had 10 or more days of symptoms and were positive despite the fact that surfaces were cleaned twice a day^1^.

Pre-symptomatic COVID-19 patients can contaminate their surroundings as well. A study of quarantined asymptomatic students rooms showed that 8/22(36%) surfaces, including linen were positive. The students later developed COVID-19^7^. This study has limitations. It is not clear which is the best method to collect the SARS-CoV-2 from MPs. Furthermore, the C_ts_ found are high and may be interpreted as having a small viral load^8^, although late amplification may have been caused by the freezing and thawing of the samples^9^,^10^.

## Conclusions

Healthcare worker’s MPs can be contaminated by SARS-CoV-2. Thus, it is possible MPs may be a part of the chain of virus transmission in healthcare settings.

## Data Availability

The data or references to data supporting the findings of this study are available within the article, further information may be requested to the corresponding author.

## ACKNOWLEDGEMENTS

We will like to thank all the health workers and scientists fighting anonymously every day during the pandemic.

